# Early immunologic response to mRNA COVID-19 vaccine in patients receiving biologics and/or immunomodulators

**DOI:** 10.1101/2021.09.11.21263211

**Authors:** Esteban Rodríguez-Martinó, Rafael Medina-Prieto, Jorge Santana-Bagur, María Santé, Petraleigh Pantoja, Ana M. Espino, Carlos A. Sariol, Esther A. Torres

## Abstract

Patients with immune conditions and immune-modifying therapies were excluded from the Covid-19 vaccine trials. Studies have shown conflicting response to different vaccines in persons receiving immune suppressors or biologics. The aim of this study is to evaluate humoral and cellular response to Covid-19 vaccines in patients with Inflammatory Bowel Disease (IBD) using biologic and/or immunomodulatory (IMM) therapies.

**Methods:** Participants are adults with IBD receiving biologics or IMM planning to receive a Covid 19 vaccine. Cellular immunity (CD4+ and CD8+ T cell levels) with flow cytometry are measured at baseline and 2 weeks after each vaccine dose. Humoral immunity (antibody titers and neutralizing capacity,VNT%) is analyzed by ELISA at baseline, 2 weeks after each dose, and 6 and 12 months after vaccine. We present the early results of the first 19 subjects. The study is approved by the IRB.

**Results:** 19 subjects (18 in biologics and 1 in IMM) who received 2 doses of the Pfizer-BioNTech vaccine are included. Total IgG antibodies increased 21.13 times after the first dose and 90 times after the second dose. VTN% increased 11.92 times after the first dose and 53.79 times after the second dose. When compared with a healthy control cohort, total IgG antibodies and VTN% were lower in the subjects after the first dose. After the second dose, IgG antibodies increased but remained lower than controls, but VTN% were similar to controls. CD4 and CD8 mean levels had an upward trend after vaccination.

**Conclusions:** Neutralizing capacity response to the vaccine in subjects was similar to a healthy cohort in spite of lower increases in total IgG antibodies. The CD4 and CD8 results observed may support the capacity to mount an effective cellular response in patients on biologics. Larger studies are needed to determine vaccine efficacy in these patients.

## Introduction

Inflammatory Bowel Diseases (IBD) – Crohn’s disease (CD) and ulcerative colitis (UC) – are characterized by chronic intestinal inflammation associated to dysregulation of the immune system. Immune-modifying agents for treatment of IBD may result in a reduced response to some vaccines [1, 2, 3]. Infliximab may be associated with suppressed CD4+ and CD8+ T-cell proliferation and activation in patients with active UC [4].

Patients with immune conditions were excluded from COVID-19 vaccine trials. Questions remain regarding the impact of medications on vaccine efficacy in this population. A study showed 100% seropositivity following two-dose Pfizer-BioNTech and NIH-Moderna COVID-19 vaccination in patients with IBD receiving biologics [5]. Infliximab has been associated with attenuated serological responses to SARS-CoV-2 when compared to gut-specific agent vedolizumab [6]. Data about antibody viral neutralization capacity (VNT%) and cellular immunity are lacking. Our aim is to evaluate humoral and cellular response to the COVID-19 vaccine in patients with IBD who are using biologic and/or immunomodulatory therapy.

## Methods

Patients with IBD between 21 and 65 years of age receiving biologics and/or immunosuppressives and planning to receive a COVID-19 vaccine were invited to participate. The study examines cellular immunity (CD4+ and CD8+ T-cell levels) via flow cytometry at baseline and 2 weeks after each vaccine dose, and humoral immunity (antibody titers and VNT%) via ELISA at baseline, 2 weeks after each dose, 6 and 12 months after completing vaccination. We report results of cellular and humoral immunity for the first 2 months in the initial subjects and compare them with a healthy cohort.

### Ethical Statement

The studies are approved by the Medical Sciences Campus IRB. Volunteers in the control group were participating in the IRB approved clinical protocol “Molecular Basis and Epidemiology of Viral infections circulating in Puerto Rico”, Pro0004333. Protocol was submitted to, and ethical approval was given by, Advarra IRB on April 21, 2020. That protocol also received ethical approval from the Medical Sciences Campus IRB.

## Results

Nineteen subjects (17 with CD and 2 with UC, 10 males) who received the BNT162b2 mRNA Pfizer-BioNTech 2-dose vaccine are included. The mean age was 34 (range 22-59). 18 participants were receiving biologic monotherapy, 1 was only on azathioprine.

Total IgG antibodies increased by 21.13 times (mean 0.715, SD 0.476, range 0.031-1.691) after the first dose and by 90.0 times after the second dose (mean 2.261, SD 0.258, range 1.66-2.58). The VNT% increased by 11.92 times after the first dose and by 53.79 after the second dose. As shown in figure 1, the total of IgG antibodies and the % of neutralizing antibodies after the first dose were significantly lower in our subjects when compared with a cohort of vaccinated healthy persons. After the second dose, total IgG antibodies increased but were still lower than healthy subjects, while the % of neutralizing antibodies was not significantly different between the 2 groups.

**Figure 1:**
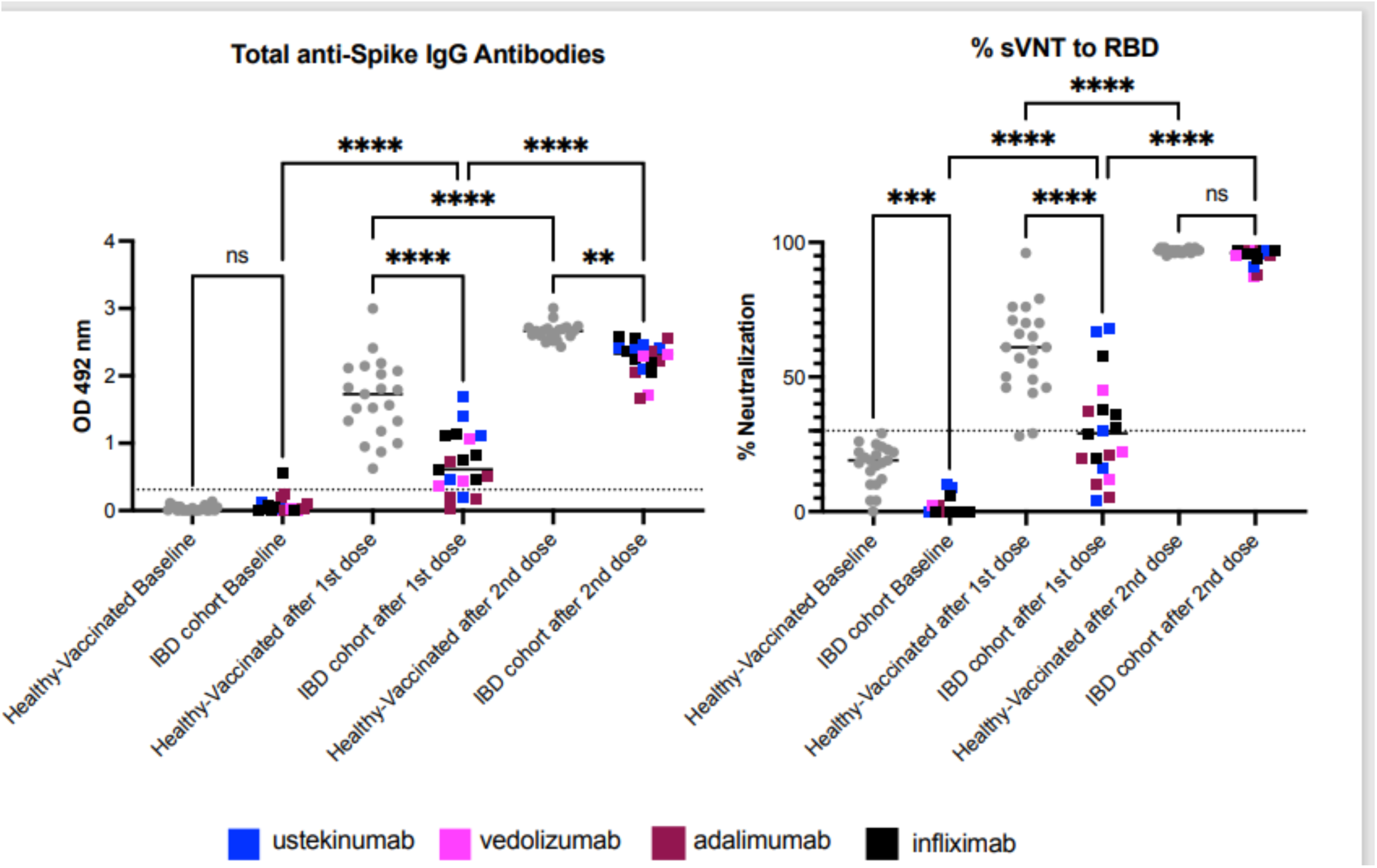
Patients with IBD on biologics show a limited humoral immune response to COVID-19 vaccine. The figure shows the results for 19 subjects with IBD compared with 21 healthy volunteers. Samples correspond to baseline (before vaccination), and 2-weeks after the first and the second Pfizer Covid-19 vaccine doses. In average samples were collected 14 days after each dose. Samples from 21 healthy volunteers were available for comparison and were collected prior to vaccination (baseline) and in average 15 to 20 days after each dose. Of the 21 healthy volunteers, eighteen (18) received Pfizer’s vaccine and three (3) Moderna’s formulation. **Panel A** shows the total anti-Spike S1 IgG antibodies. Results show that one vaccine dose in IBD patients triggers a limited antibodies response which is significantly lower compared to the healthy cohort. Five patients with IBD had no detectable antibodies after the first dose. After the second dose all IBD patients had detectable antibodies, however they were still significantly lower compared to the healthy control group. Panel B shows the antibodies’ blocking capabilities measured by a surrogate viral neutralization assay (sVNT) and results are provided as percentage of neutralization. After the first vaccine dose, only 38% of IBD patients developed neutralizing activity (n=8) compared with 90.4% of the healthy individuals (n=19). Three (3) patients with IBD developed borderline neutralizing activity and 47.6% (n=10) did not develop neutralizing activity. The second vaccine shot boosted the neutralizing activity and 100% of the patients had detectable neutralizing activity with a magnitude similar to the healthy cohort. Interestingly, and as a potential consequence of the immunosuppression therapy, the blocking baseline activity of the healthy individuals is significantly higher than the IBD group. However, all those values were below the detection threshold. The threshold for the total antibodies was 0.312 and for the blocking activity was 30%. Statistical significance was determined by One-way ANOVA multiple comparisons to test for increase or decrease among samples. Tukey’s multiple comparisons test was performed as post-hoc test *p*<0.05 was considered significant.

Mean CD4 levels were 809.95± 234.69, 818.63 ±238.94, and 967.16 ± 396.72 for visits 1, 2, and 3, respectively. Mean CD8 levels were 361.00± 141.34, 416.26 ± 202.98 and 470.11± 284.70 for visits 1, 2, and 3, respectively. Both CD4 and CD8 mean levels showed an upward trend after vaccination. (Fig 2).

**Figure 2.**
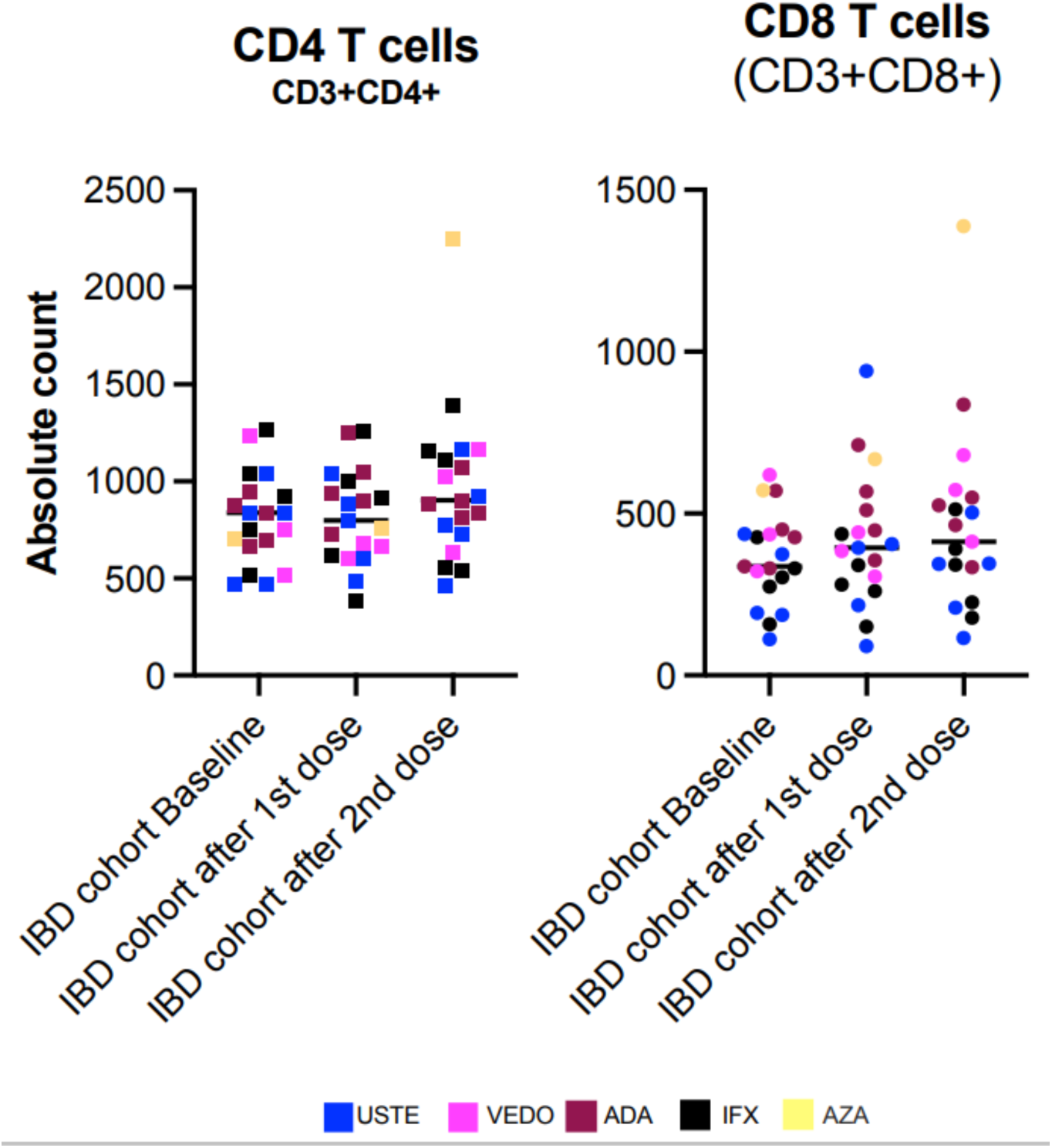
Vaccine response of CD4 and CD8 T-cells in subjects with IBD. **Panel A** shows the individual absolute CD4 count at baseline and after the first and second dose of the vaccine, identified with the specific biologic being used. Five subjects were on infliximab, 5 on adalimumab, 5 on ustekinumab, 3 on vedolizumab and 1 on azathioprine. There is a mild increase in mean CD4 count after the second vaccine dose. **Panel B** shows the same parameters for the individual total CD8 counts. There is a small progressive increase in mean CD8 counts after each vaccine dose. These findings suggest the vaccine is safe and do not support a negative effect of vaccination in the cellular response of subjects with IBD receiving biologic/immunomodulatory therapy. Statistical significance was determined by One-way ANOVA multiple comparisons to test for increase or decrease among samples. Tukey’s multiple comparisons test was performed as post-hoc test *p*<0.05 was considered significant. IBD= Inflammatory Bowel Disease, USTE = ustekinumab, VEDO= vedolizumab, ADA = adalimumab, IFX = infliximab, AZA = azathioprine.

## Discussion

To our knowledge, this is the first report of both serological and cell-mediated components in response to COVID-19 vaccines in patients receiving biologics for IBD. Although the antibody response was low, neutralizing capacity after the second dose was similar to that of a healthy cohort. The preservation and increase of the CD4+ and CD8+ T-cells after each dose indicates that patients on biologics for IBD are potentially capable of mounting a quantitative effective cell-mediated immune response. Whether this cellular and humoral response is protective or may require additional vaccine boosting remains unknown at present. Limitations of the study include the small size of the population and the short follow-up period. Larger studies with longer follow up and comparison between biologics with different mechanisms of action are necessary to establish the efficacy of the vaccine in these patients.

## Data Availability

All data related to this work is available upon request

## Author Contributions

Drs. Rodríguez, Santana, Torres and Sariol had full access to all the data in the study and take responsibility for the integrity of the data and the accuracy of the data analysis. Concept and design: Rodríguez-Martinó, Santana, Sariol, Torres, Santé. Acquisition, analysis and interpretation of data: all authors. Drafting of the manuscript: Rodríguez-Martinó, Sariol, Torres. Critical revision of the manuscript for important intellectual content: all authors. Statistical analysis: Sariol, Pantoja. Obtained funding: Torres, Sariol, Espino. Administrative, technical, or material support: Sariol, Santé, Pantoja. Supervision: Torres, Sariol.

## Additional contributions

We acknowledge the contributions of Sofia Ojeda BS (UPR School of Medicine) with subject recruitment and data collection. No financial compensation was received.

